# Sex-Specific Effect of MTSS1 Downregulation on Dilated Cardiomyopathy

**DOI:** 10.1101/2024.02.28.24303451

**Authors:** Dongwook Choe, Megan Burke, Jeffrey A Brandimarto, Ingrid Marti-Pamies, Jaime Yob, Yifan Yang, Michael P Morley, Theodore G. Drivas, Sharlene Day, Scott Damrauer, Xiao Wang, Thomas P Cappola

**Affiliations:** Division of Cardiovascular Medicine, all at the University of Pennsylvania Perelman School of Medline, Philadelphia, PA, USA; Penn Cardiovascular Institute, all at the University of Pennsylvania Perelman School of Medline, Philadelphia, PA, USA; Division of Translational Medicine and Human Genetics, all at the University of Pennsylvania Perelman School of Medline, Philadelphia, PA, USA; Deparment of Genetics, all at the University of Pennsylvania Perelman School of Medline, Philadelphia, PA, USA

## Abstract

MTSS1 (metastasis suppressor 1) is an I-BAR protein that regulates cytoskeleton dynamics through interactions with actin, Rac, and actin-associated proteins. In a prior study, we identified genetic variants in a cardiac-specific enhancer upstream of *MTSS1* that reduce human left ventricular (LV) MTSS1 expression and associate with protection against dilated cardiomyopathy (DCM). We sought to probe these effects further using population genomics and *in vivo* murine models. We crossed *Mtss1*^*-/-*^ mice with a transgenic (*Tg*) murine model of human DCM caused by the D230N pathogenic mutation in *Tpm1* (tropomyosin 1). In females, *Tg/Mtss1*^+/-^ mice had significantly increased LV ejection fraction and reduced LV volumes relative to their *Tg/Mtss1*^+/+^ counterparts, signifying partial rescue of DCM due to *Mtss1* haploinsufficiency. No differences were observed in males. To study effects in humans, we fine-mapped the *MTSS1* locus with 82 cardiac magnetic resonance (CMR) traits in 22,381 UK Biobank participants. *MTSS1* enhancer variants showed interaction with biological sex in their associations with several CMR traits. After stratification by biological sex, associations with CMR traits and colocalization with *MTSS1* expression in the Genotype-Tissue Expression (GTEx) Project were observed principally in women and were substantially weaker in men. These findings suggest sex dimorphism in the effects of MTSS1-lowering alleles, and parallel the increased LV ejection fraction and reduced LV volumes observed female *Tg/Mtss1*^+/-^ mice. Together, our findings at the *MTSS1* locus suggest a genetic basis for sex dimorphism in cardiac remodeling and motivate sex-specific study of common variants associated with cardiac traits and disease.

---

MTSS1 (metastasis suppressor 1) is an I-BAR protein that regulates cytoskeleton dynamics through interactions with actin, Rac, and actin-associated proteins.^1^ MTSS1 is also known to influence cellular motility and adhesion and has been implicated in development of myocardial trabeculation.^2^ In a prior study, we identified genetic variants in a cardiac-specific enhancer upstream of *MTSS1* that reduce human left ventricular (LV) MTSS1 expression and associate with protection against dilated cardiomyopathy.^3^ We sought to probe these effects further using population genomics and *in vivo* murine models. All studies were conducted in accordance with institutional ethical approvals.

We identified 10,895 community-dwelling men (mean±SD age 55.8±7.5 years) and 11,486 women (54.4±7.2 years) in the UK Biobank (project 65965) who underwent genotyping and cardiac magnetic resonance (CMR) imaging. We fine-mapped associations between 82 CMR traits and variants within 100kb of the *MTSS1* enhancer at chr 8:124845117 and tested for colocalization with regulation of LV *MTSS1* expression in 386 subjects in the Genotype-Tissue Expression Project (GTEx). There were strong colocalizations among *MTSS1* enhancer variants, CMR traits, and *MTSS1* gene expression (**Figure, panel A)**. MTSS1-lowering alleles were linked to smaller LV volumes and higher ejection fraction (ColocPr: LV end-systolic volume [LVESV]=0.993, LV end-diastolic volume [LVEDV]=0.764, LV ejection fraction [LVEF]=0.994) and to enhanced contractile function measured by global LV strain (ColocPr: Ecc_Global = 0.995) and by multiple measures of regional LV strain.

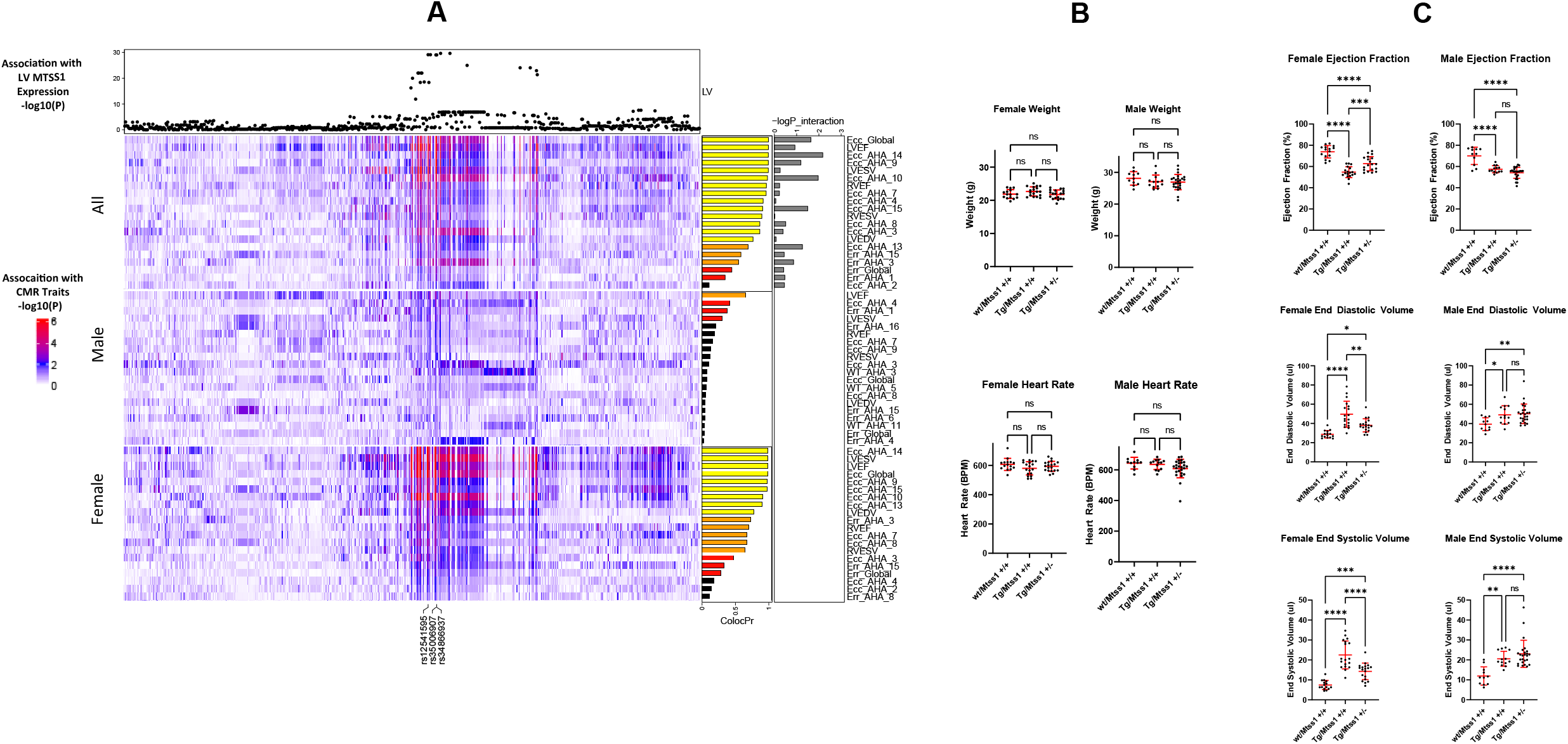
**A:** Association and colocalization of *MTSS1* enhancer variants with LV MTSS1 expression in GTEx and CMR traits in UK Biobank. The abscissa displays the region within 100 KB of the enhancer, which is centered at chr 8:124845117 and marked by three previously published variants (rs12541595, rs35006907, rs3466937). Associations with MTSS1 expression are displayed as -log_10_P-values on the top row. Associations with 20 of 82 CMR traits were identified and are displayed in three separate heatmaps comprising all patients, male only, and female only; traits are labeled using standard abbreviations for ventricular volumes, ejection fraction, and global/regional LV strain. Colocalization probabilities are indicated in barplots to the right. Interaction between biological sex and lead MTSS1 variants in their associations with CMR traits are displayed as -logP_interaction. **B:** Total body weight and heart rate for male (*wt/Mtss1*^+/+^, n=11; *Tg/Mtss1*^+/+^, n =13; *Tg/Mtss1*^+/-^, n= 25) and female (*wt/Mtss1*^+/+^, n=15; *Tg/Mtss1*^+/+^, n=8; *Tg/Mtss1*^+/-^ *Mtss1*^+/-^, n=19) mice. **C:** Ejection fraction, LV end diastolic volume, and LV end systolic volume of mice from (B) by 2D echocardiography. *P < 0.05, **P < 0.01, ***P < 0.001, **** P < 0.0001 by one-way ANOVA with Tukey post-hoc test.

We next used transgenic murine models to determine whether MTSS1 reduction could rescue dilated cardiomyopathy. The *TPM1* D230N variant is known to cause progressive dilated cardiomyopathy, and the *Tpm1* D230N transgenic mouse model (*Myh6-Tpm1*D230N*) similarly has reduced LVEF and increased LVEDV/LVESV by two months of age.^4^ We crossed mice harboring this transgene (*Tg*) with *Mtss1* knockout mice *(Mtss1*^*-/-*^)^3^ to generate *wt/Mtss1*^*+/+*^, *Tg/Mtss1*^*+/+*^ and *Tg/Mtss1*^*+/-*^ strains. At 5 months of age we assessed these mice with two-dimensional echocardiography, blinded to genotype and stratified by biological sex. No statistically significant differences were detected for body weight or heart rate across the genotype groups **(Figure, panel B)**. Statistically significant LVEF reductions and LVESV/LVEDV increases were observed for *Tg/Mtss1*^+/+^ mice relative to their wild-type controls regardless of biological sex, consistent with development of dilated cardiomyopathy **(Figure panel C)**. For female mice expressing the *Tpm1* D230N transgene, we observed that *Tg/Mtss1*^+/-^ mice had significantly increased LVEF and reduced LVESV/LVEDV relative to their *Tg/Mtss1*^+/+^ counterparts **(Figure panel C)**. These findings signify partial rescue of dilated cardiomyopathy in female mice by reducing MTSS1. In contrast, no differences were observed between *Tg/Mtss1*^+/-^ and *Tg/Mtss1*^+/+^ male transgenic mice **(Figure, panel C)**. Control experiments indicated no difference in transgene expression by sex. These findings reveal sex-dimorphism in the effects of Mtss1 reduction, with a partial rescue of dilated cardiomyopathy limited to females.

We returned to the CMR data to explore whether sex-specific effects were also present in humans. The lead *MTSS1* enhancer variants showed interaction with biological sex in their associations with several CMR traits (**Figure, panel A**). After stratification by biological sex (**Figure, panel A)**, associations with CMR traits and colocalization with *MTSS1* expression were observed principally in women (female ColocPr: LVESV=0.99, LVEDV=0.79, LVEF = 0.99, Ecc_Global” = 0.99) and were substantially weaker in men (male ColocPr: LVESV=0.31, LVEDV= 0.05, LVEF = 0.67, “Ecc_Global” = 0.09). These findings suggest sex dimorphism in effects of MTSS1-lowering alleles, and parallel the increased LVEF and reduced LVESV/LVEDV observed female *Tg/Mtss1*^+/-^ mice. Thus, the observed sex-specific differences in mice are highly concordant with the sex-specific human genetic findings.

Differences in presentation and prognosis of heart disease associated with biological sex have been studied for decades. Here, we report that MTSS1 may have a cardioprotective effect against dilated cardiomyopathy for women but not for men, suggesting a biological basis for sex dimorphism in cardiac remodeling. Discerning the specific underlying mechanisms will require additional experiments, including use of gonadectomized models to test for hormonally mediated effects. Likewise, UKBB is a healthy population sample of European ancestry; additional work is required to quantify sex-specific genetic effects in cohorts of cardiomyopathy patients, in diverse ancestries, and in heart failure with preserved ejection fraction, which has higher prevalence in women. Our findings suggest common genetic variants linked to cardiac traits and disease should be probed for difference by sex to assess risk more accurately and to identify sex-specific mechanisms of disease.

## Data Availability

All data produced in the present study are available upon reasonable request to the authors

## Acknowledgements

We are grateful to Dr. Jil Tardiff for providing breeding pairs of *Tpm1* D230N transgenic mice.

